# The effectiveness of gender-based violence prevention among adolescents aged 10 to 19 years in Southeast Asia: A systematic review

**DOI:** 10.1101/2025.02.17.25322431

**Authors:** Hoa H. Nguyen, Kien G. To, Van TH. Hoang, Lu Gram, Kathryn M. Yount

## Abstract

**Background:** Gender-based violence (GBV) is a major public health problem with wide-ranging negative physical and mental health consequences for survivors. The global economic costs of GBV are estimated at US$1.5 trillion. Adolescence is a period of high risk for GBV. The evidence-based GBV prevention programs among adolescents in Southeast Asia are not well understood. This paper synthesizes GBV prevention intervention studies in this region’s adolescents aged 10-19 years.

**Methods:** A systematic review was conducted following the PRISMA guidelines. PubMed, Medline, Embase, Web of Science, and PsycINFO were used to search for potential articles published until 3^rd^ September 2024. We screened identified articles following consistent inclusion and exclusion criteria. We assessed the risk of bias using the ROBINS-I tool for observational and quasi-experimental studies and the RoB 2 tool for randomized controlled trials.

**Results:** Eight of 1689 identified articles were included in the review (five quasi-experimental designs, two pre-test and post-test designs, and one cluster randomized controlled trial). Most studies (7/8) focused on school-based educational programs. The risk of bias in the cluster randomized controlled trial was “some concern” level, compared with the moderate to serious level of the non-randomized studies. One pre-and post-test study directly measured GBV behaviour and found that children reported fewer experiences of physical assault (mean difference: 1.1; 95% CI: 0.6, 1.6) and psychological aggression (1.5; 0.8, 2.3) after the intervention. The remaining studies evaluated the knowledge, attitudes, and skills of participants as primary outcomes in sexual violence prevention programs. Three of four studies that assessed changes in knowledge showed significant improvements. Inconsistent findings were found in association with attitudes and skills of GBV intervention prevention programs.

**Conclusions:** GBV intervention studies among adolescents in Southeast Asia are rare and evidence of effectiveness is generally weak. Rigorous RCTs that assess program impacts on GBV behaviour, as well as knowledge, attitudes, and skills, are needed. Programs that incorporate classroom activities and online learning with Facebook/Zalo/Instagram assignments and games may show promise for rigorous evaluation in Southeast Asia.

**Registration:** This systematic review was registered on PROSPERO (CRD42023476059).

## Background

Gender-based violence (GBV) is a major public health problem (1). GBV includes physical, sexual, verbal, emotional, and economic forms of dating or intimate partner violence, online or in-person forms of stalking, non-partner sexual violence, and sexual harassment (1). Exposure to GBV has negative consequences for physical and mental health, including but not limited to injury, unintended pregnancy, HIV, sexually transmitted infections, post-traumatic stress disorder, depression, or death (2, 3). The global economic costs of GBV are estimated to be about US$1.5 trillion, or 2% of global gross domestic product (GDP) (4).

Adolescence is a period of heightened risk for GBV (2, 3). Globally, the lifetime prevalence of intimate partner violence (IPV) among adolescent girls (15-19 years) was 24% (5). In Southeast Asian countries (Malaysia, Myanmar, Brunei, Thailand, Vietnam, Cambodia, East Timor, Indonesia, Laos, the Philippines and Singapore), the lifetime prevalence of sexual victimization among those aged 12 to 21 years was 3%-65% for girls and 3%-42% for boys (6). Of 33,184 adolescents across Thailand, Timor-Leste, Laos, Indonesia and the Philippines (mean age: 14.6 years), 30.6% of participants experienced any past month bullying victimization (boys: 33.9%, girls: 27.5%) (7). The physical, social aggression, and verbal violence prevalence among 712 Vietnamese secondary school students was 8.4%, 31.2%, and 11.9% (8).

To date, school-based GBV prevention programs among adolescents are limited to low-and middle-income settings. A current meta-analysis found that school-based interventions may be effective in reducing victimization and perpetration of GBV; however, all effects were highly imprecise and findings for GBV outcomes were inconsistent (9). Most intervention studies also have been conducted in North America, Europe, Africa, and South America, despite high prevalence rates of GBV in Southeast Asia, as well. We aimed to fill this evidence gap by conducting a systematic review of GBV prevention studies among adolescents in Southeast Asia. Findings can inform recommendations for future rigorous research to develop and test more widely suitable school-based intervention programs to reduce the prevalence of GBV in Southeast Asia, with potential regional impacts on adolescent health.

## Methods

The National Institute for Health Research of the University of York approved the protocol registration for a systematic review (PROSPERO ID number: CRD42023476059) (10). We used the Preferred Reporting Items for Systematic Reviews and Meta-Analyses (PRISMA) tool for our systematic review checklist (11).

### Searches

We used five databases (PubMed, Embase, Web of Science, Medline, and PsycINFO) to search for potential articles on 6^th^ January 2024, and we updated the search on 3^rd^ September 2024. We developed, piloted, and refined the search string for each database, using the MeSH terms and text words (12). These words related to Southeast Asia, intervention studies, adolescents, and gender-based violence. The search strategy we used in the Embase database is presented in **Table 1**, and the search strings used with other databases are available in **Supplemental File 1**.

**Table 1:**
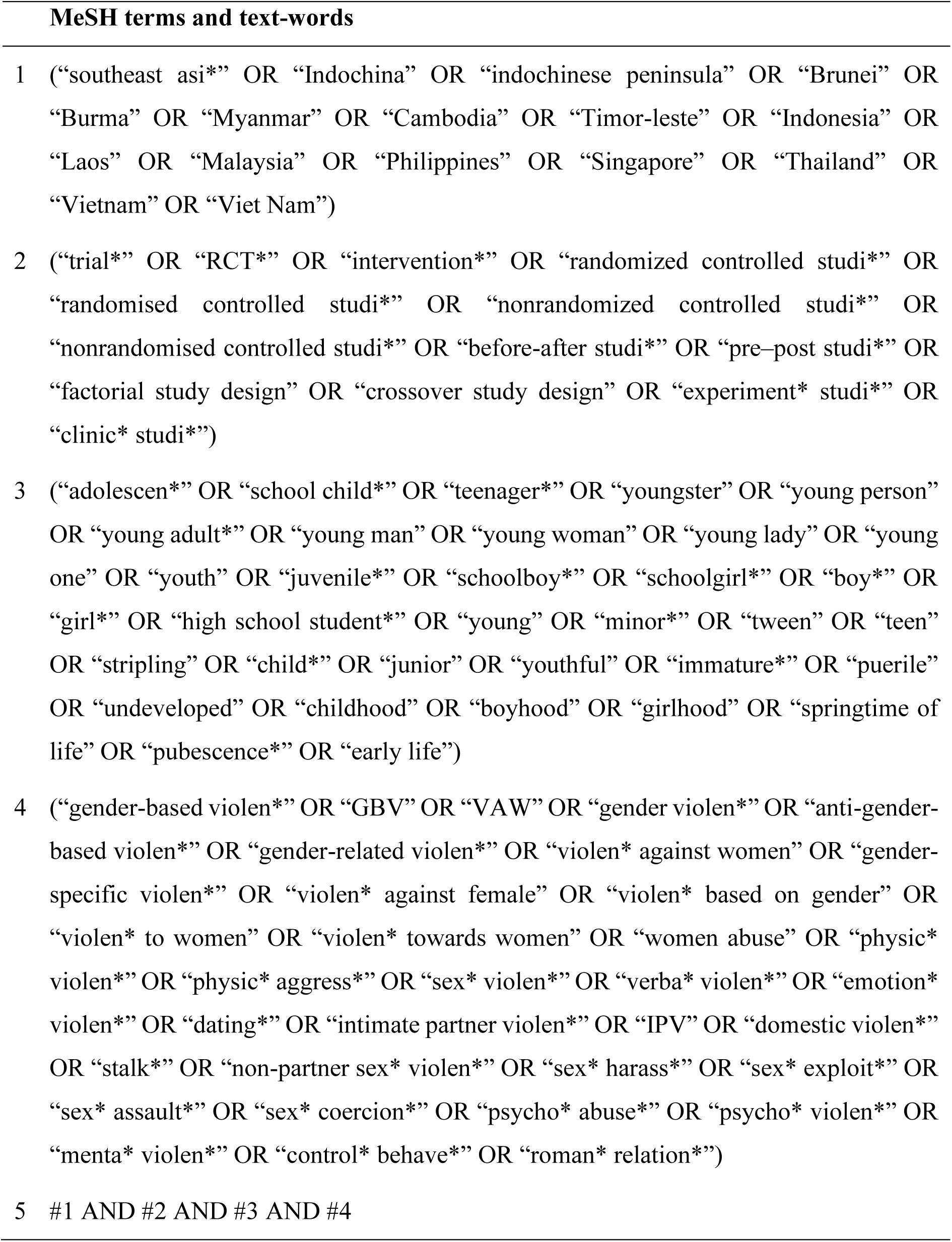
Search strategy in the Embase database.

| MeSH terms and text-words |  |
| --- | --- |
| 1 | ("southeast asi*" OR "Indochina" OR "indochinese peninsula" OR "Brunei" OR "Burma" OR "Myanmar" OR "Cambodia" OR "Timor-leste" OR "Indonesia" OR "Laos" OR "Malaysia" OR "Philippines" OR "Singapore" OR "Thailand" OR "Vietnam" OR "Viet Nam") |
| 2 | ("trial*" OR "RCT*" OR "intervention*" OR "randomized controlled studi*" OR "randomised controlled studi*" OR "nonrandomized controlled studi*" OR "nonrandomised controlled studi*" OR "before-after studi*" OR "pre-post studi*" OR "factorial study design" OR "crossover study design" OR "experiment* studi*" OR "clinic* studi*") |
| 3 | ("adolescen*" OR "school child*" OR "teenager*" OR "youngster" OR "young person" OR "young adult*" OR "young man" OR "young woman" OR "young lady" OR "young one" OR "youth" OR "juvenile*" OR "schoolboy*" OR "schoolgirl*" OR "boy*" OR "girl*" OR "high school student*" OR "young" OR "minor*" OR "tween" OR "teen" OR "stripling" OR "child*" OR "junior" OR "youthful" OR "immature*" OR "puerile" OR "undeveloped" OR "childhood" OR "boyhood" OR "girlhood" OR "springtime of life" OR "pubescence*" OR "early life") |
| 4 | ("gender-based violen*" OR "GBV" OR "VAW" OR "gender violen*" OR "anti-gender-based violen*" OR "gender-related violen*" OR "violen* against women" OR "gender-specific violen*" OR "violen* against female" OR "violen* based on gender" OR "violen* to women" OR "violen* towards women" OR "women abuse" OR "physic* violen*" OR "physic* aggress*" OR "sex* violen*" OR "verba* violen*" OR "emotion* violen*" OR "dating*" OR "intimate partner violen*" OR "IPV" OR "domestic violen*" OR "stalk*" OR "non-partner sex* violen*" OR "sex* harass*" OR "sex* exploit*" OR "sex* assault*" OR "sex* coercion*" OR "psycho* abuse*" OR "psycho* violen*" OR "menta* violen*" OR "control* behave*" OR "roman* relation*") |
| 5 | #1 AND #2 AND #3 AND #4 |

### Study Inclusion and Exclusion Criteria

The study inclusion and exclusion criteria of this systematic review are presented in **Table 2** for more detail. After finalizing the eligible articles from five databases, we emailed the corresponding authors to find additional articles if applicable. We also checked the reference list of these articles to search for other relevant studies. However, there were no additional studies for our systematic review.

**Table 2:** Study inclusion and exclusion criteria.

| Inclusion criteria | Exclusion criteria |
| --- | --- |
| We included any quantitative study reporting on GBV intervention programs among adolescents in Southeast Asia. The screening of study titles and abstracts and the full-text review were based on specific criteria below:<br>✓ Population: Adolescents aged 10 to 19 years<br>✓ Intervention: Any intervention programs<br>✓ Outcome: e.g., Violence prevalence/incidence<br>✓ Publication type: English and Vietnamese articles<br>✓ Research type: Human subject<br>✓ Date: Any time<br>✓ Countries: Any countries in Southeast Asia based on World Bank country classification * | We excluded any publication that did not meet these inclusion criteria and/or was a duplicate of an included study, a study protocol, a narrative review, a commentary, or an editorial letter. |

\* World Bank Group. ASEAN countries. 2024.

### Study Quality Assessment

Two authors assessed the risk of bias in the included studies, using the ROBINS-I and the RoB 2 tools provided in **Supplemental File 2** (13). RoB 2 was used to assess the risk of bias in cluster randomized control trials. This tool has five domains: the randomization process, deviations from the intended interventions, missing outcome data, measurement of the outcome, and selection of the reported result. Each domain has three options: low risk (+), some concerns (!), and high risk (-). The overall risk of bias measurement of RoB 2 was assessed depending on the highest-level result of these domains. ROBIN-I was used to assess the risk of bias in non-randomized control trials. It has seven bias domains relating to confounding, selection of participants, classification of intervention, deviation from intended intervention, missing data, measurement of outcomes, and selection of the reported results. Each part has five levels: low (+), moderate (-), serious (x), critical (!), and no information. The overall risk of bias measurement of ROBIN-I was assessed depending on the highest-level result of these parts.

### Data Extraction Strategy

Two researchers independently screened titles, abstracts and full text, using the above-mentioned inclusion and exclusion criteria. We reviewed and confirmed the final screening results and resolved disagreements by consensus. A third person resolved any outstanding conflicts. Two authors reviewed and confirmed the extracted data for the following variables: Study characteristics (author, year of publication, country, study design, sample size), participants’ characteristics (age, sex, eligible criteria for participants’ recruitment), type and measurement of violence, intervention programs, data collection tools, and study results (effect sizes, 95% confidence interval (CI), and p-values).

### Data Synthesis and Presentation

We did not conduct a meta-analysis or any sub-group analyses due to the small number of studies that met the inclusion criteria, high degree of heterogeneity in study outcomes, participant characteristics, and intervention designs across included studies. Instead, we conducted a narrative analysis of evidence for all GBV outcomes related to the included intervention programs. We synthesized data and presented them in tables, figures, and supplements. In the main text, we reported the flowchart of the included studies, study characteristics, and findings of the included studies. In the supplement files, we summarized the search strategies, results of risk of bias, and the PRISMA checklist for our systematic review.

## Results

### Review Statistics

#### Results of the search process

Figure 1 summarizes the results of the search process. There were 1086/1689 potential articles found from five databases after excluding duplication. We screened titles, abstracts and full texts to finalize eight eligible articles. We contacted the corresponding authors of these articles and searched the reference lists to find more research. However, there was no more eligible research for our synthesis. Ultimately, eight out of 1689 articles were included in the systematic review (Figure 1) (14–21).

**Figure 1:**
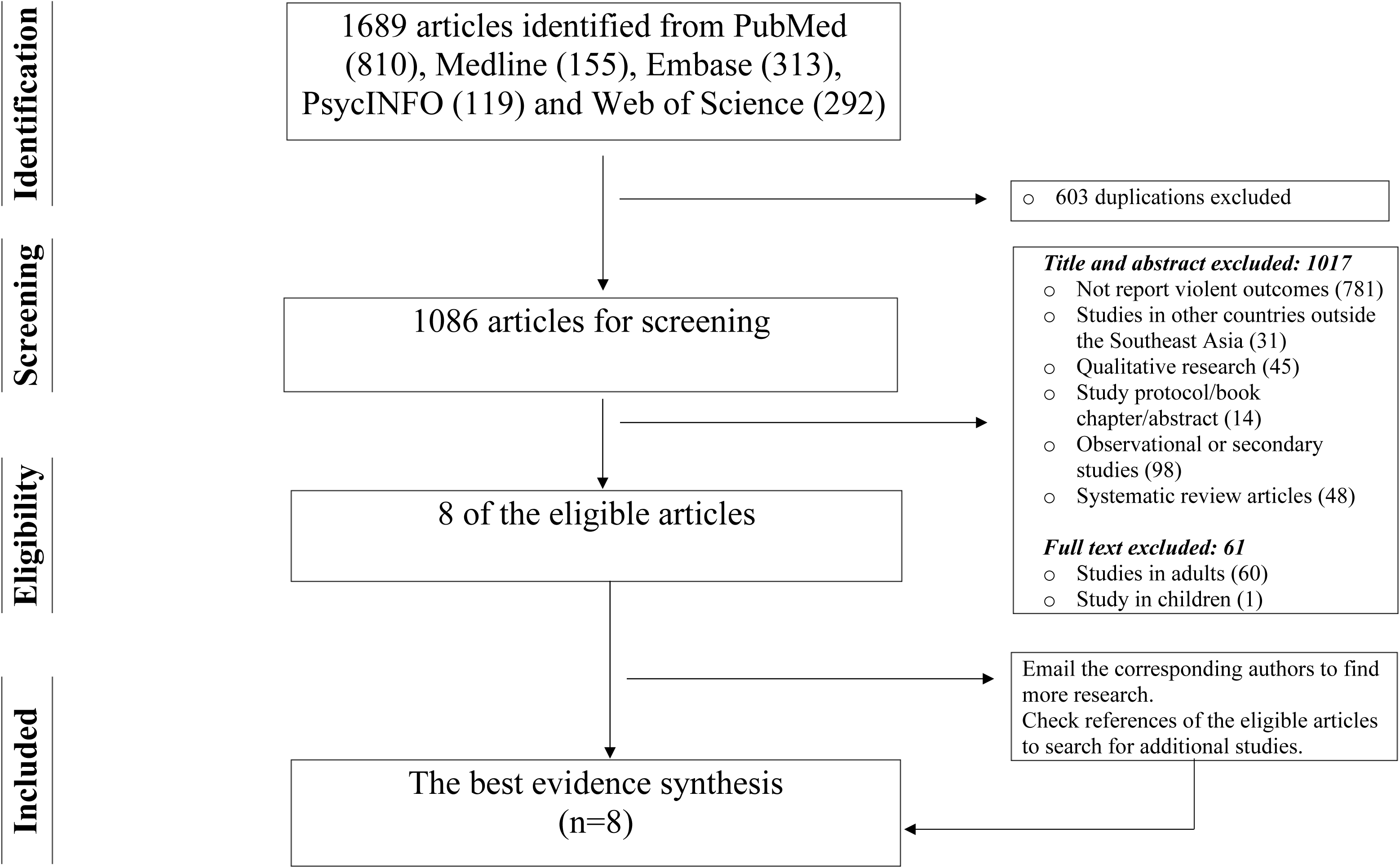
Prisma flowchart of the included studies

#### Characteristics of included studies

Study characteristics are summarized in **Table 3**. Of eight included studies, five were quasi-experimental studies (14, 15, 17, 18, 21), two were pre-and post-test research (16, 20), and one was a cluster randomized controlled trial (19). The sample size ranged from 37 (16) to 864 (15) across studies, and 6 of 8 studies had more female than male participants (15, 17–21). Half of the studies (4 of 8) were conducted in Indonesia (15, 17, 19, 21), and the remainder were conducted in Thailand (14, 18), Bali (16), and Philippines (20).

**Table 3:**
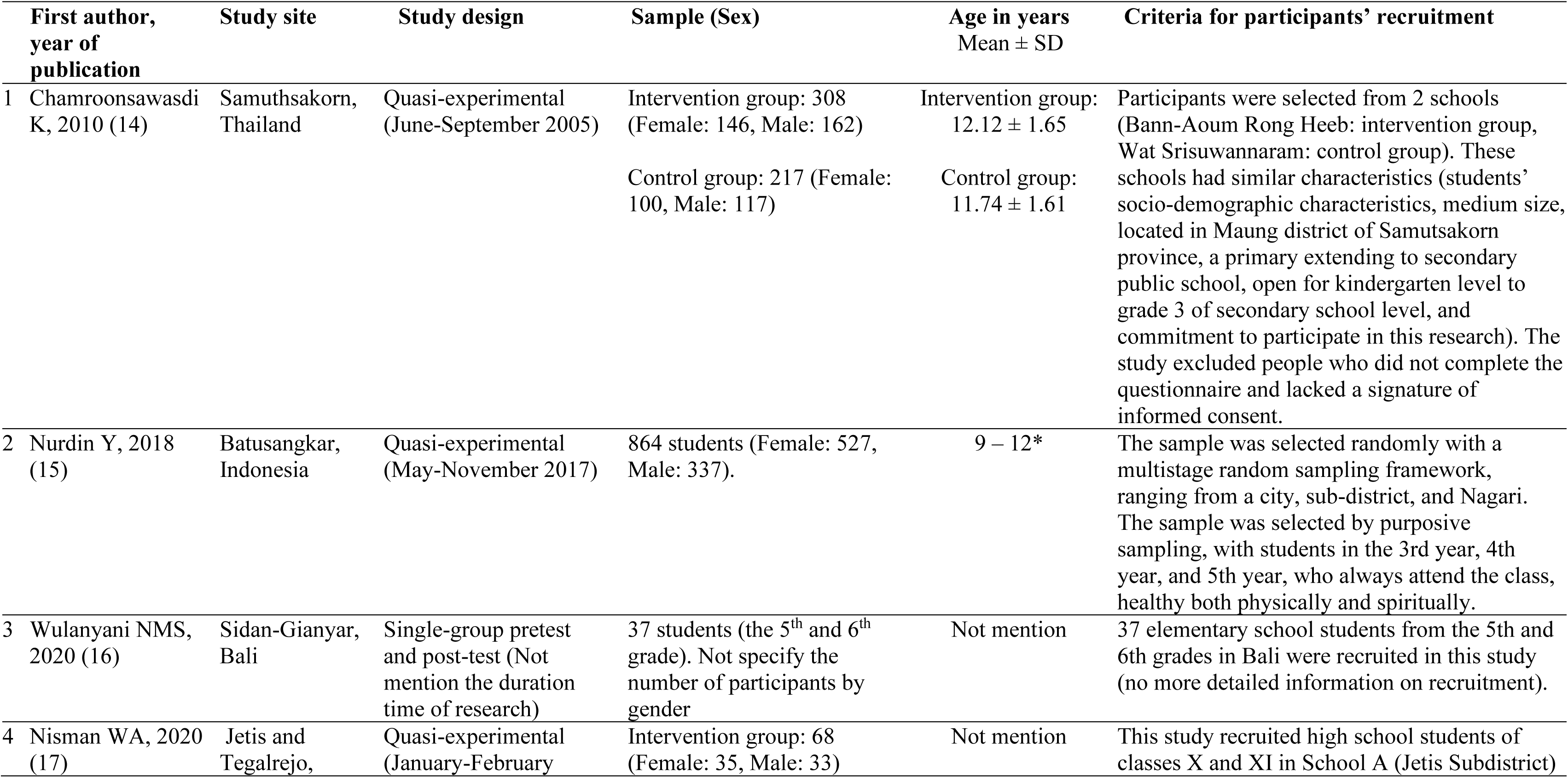
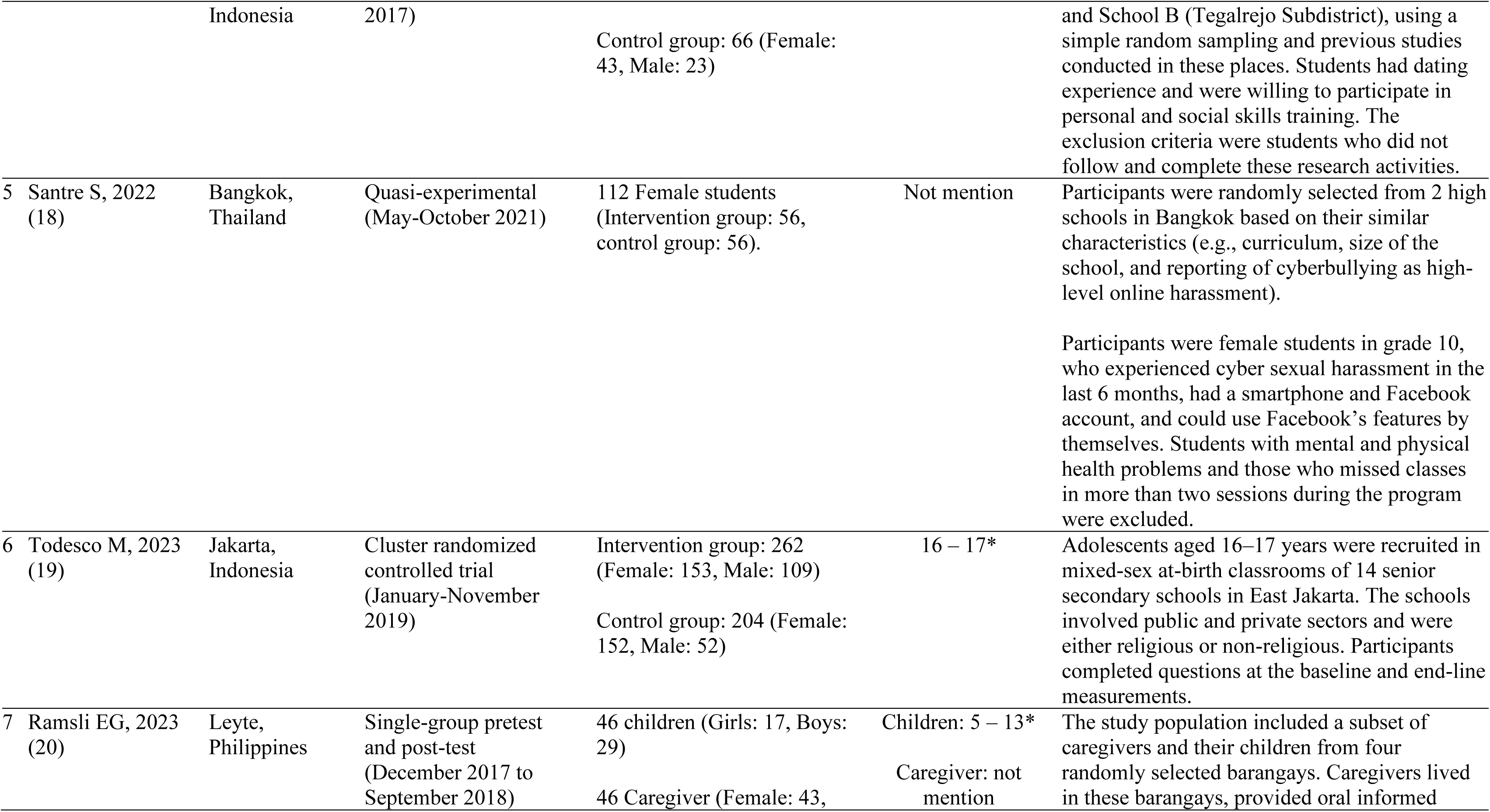
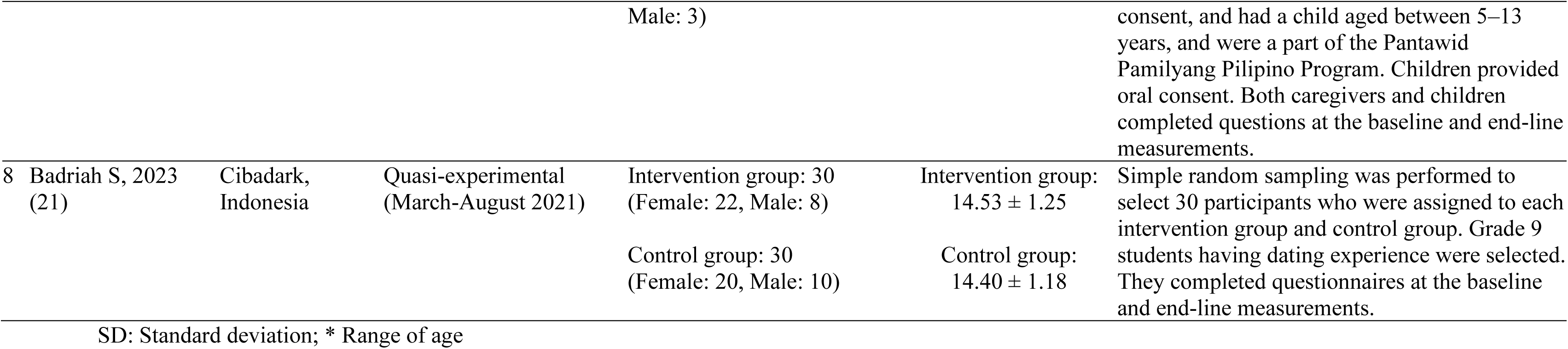
Characteristics of included studies.

#### Characteristics of GBV intervention programs

**Table 4** describes the intervention programs, data collection tools, and the findings of the included studies. Most studies (7 of 8) focused on school-based educational programs (14–19, 21). The intervention time per session varied from 45 minutes (19) to 150 minutes (14). Many intervention/experimental groups received various programs (e.g., presentations, story discussions, videos, roleplay, leaflets, scenarios, poster images, books, or questions). Meanwhile, most control groups did not receive any programs. For the outcomes assessed, one study directly measured physical assault and psychological aggression (20). Tools for data collection varied considerably, including a self-administered questionnaire (14, 18, 19), validated instruments (17, 20, 21), an animated video and game (16), or the Neherta model (15). Knowledge, attitudes, or skills toward sexual violence prevention were mentioned in six out of eight studies (14–18, 21).

**Table 4:**
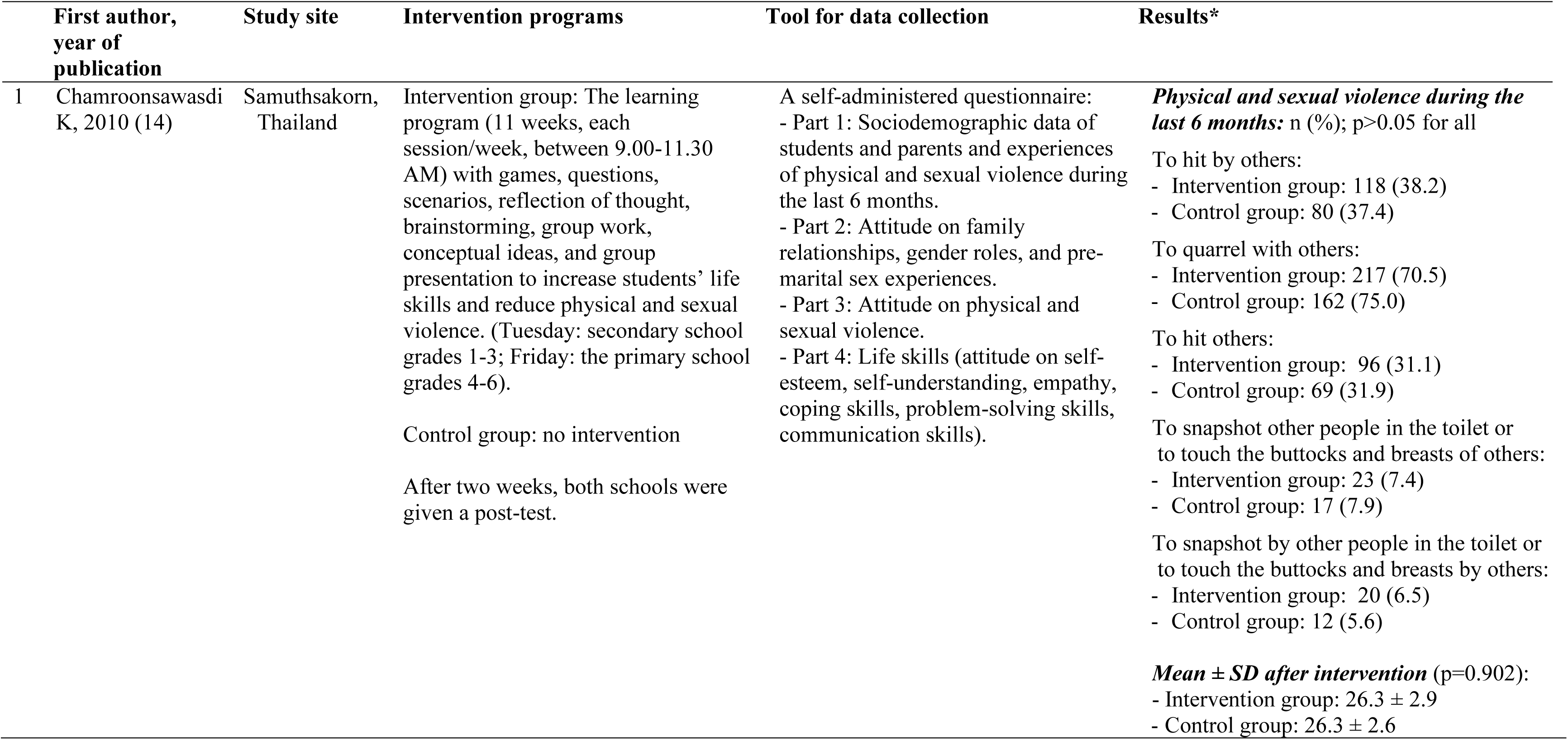
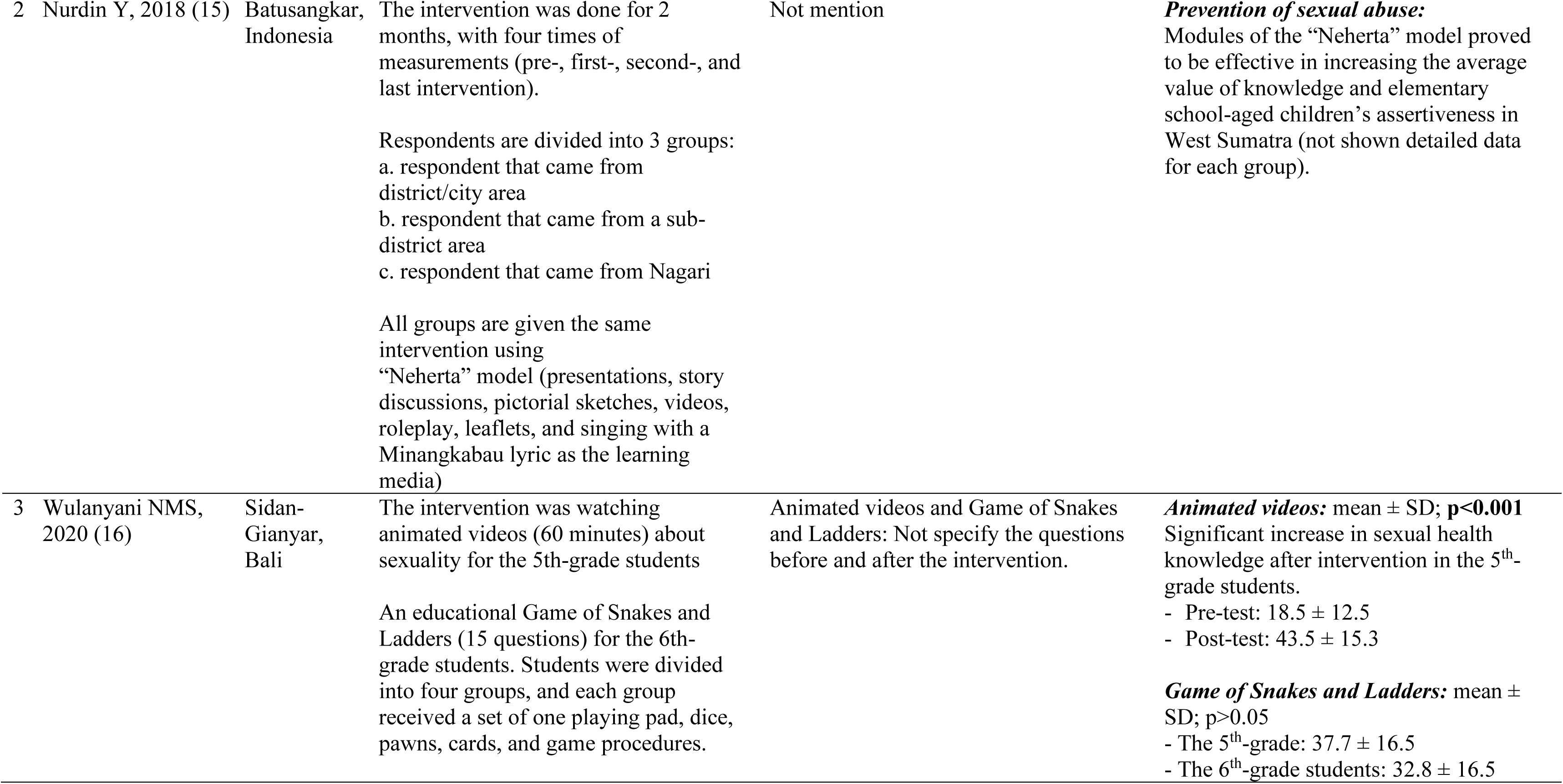
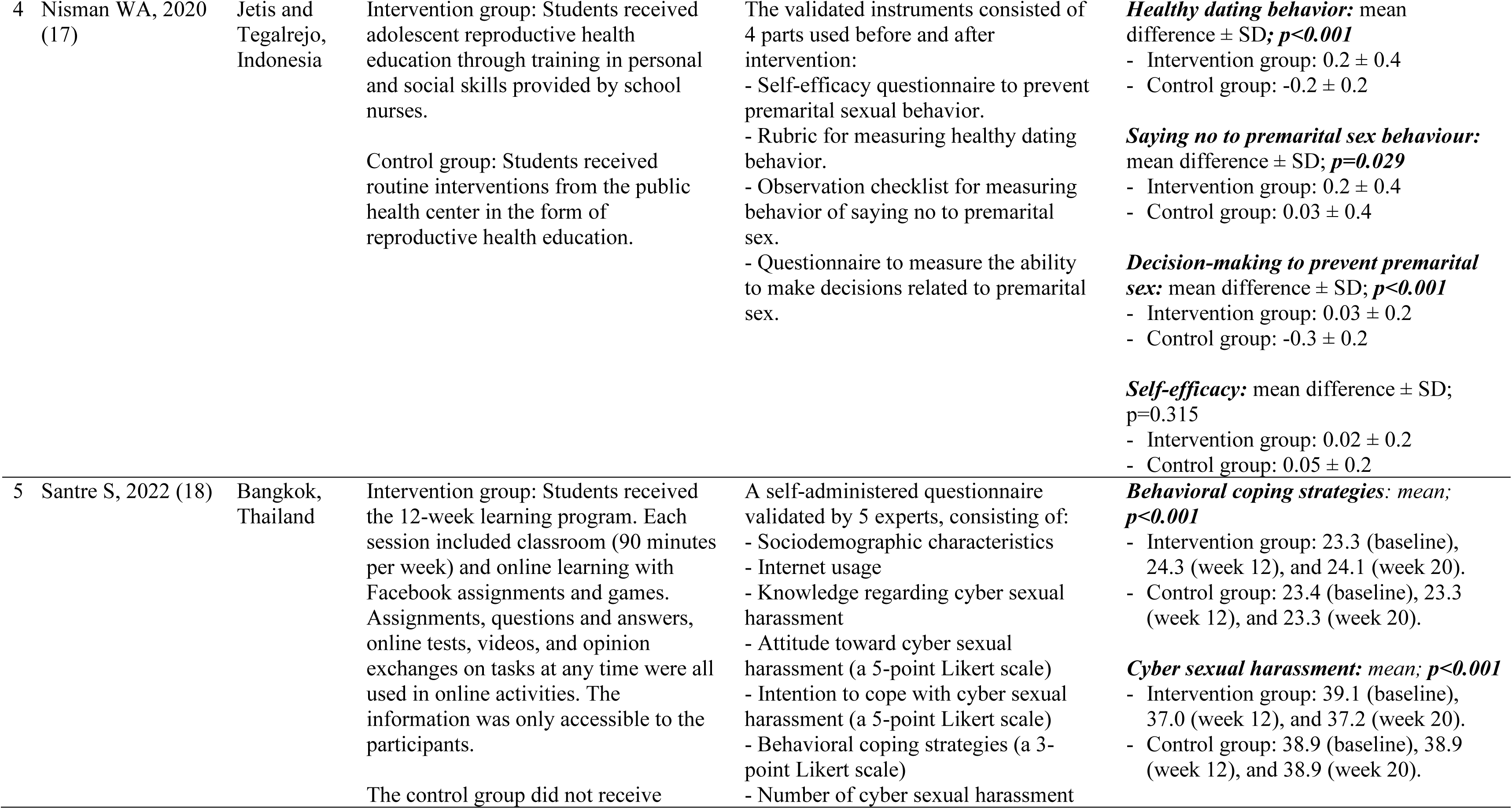
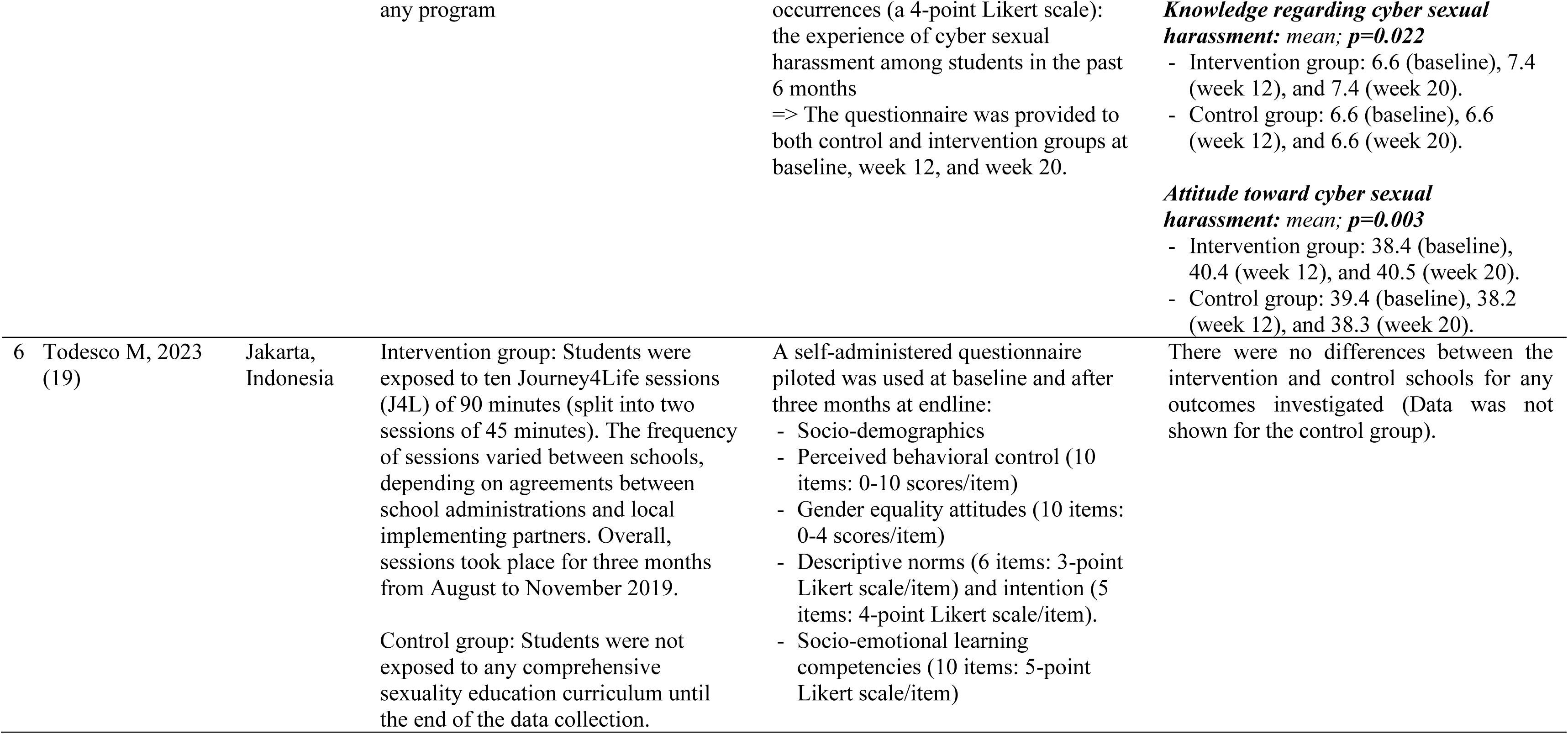
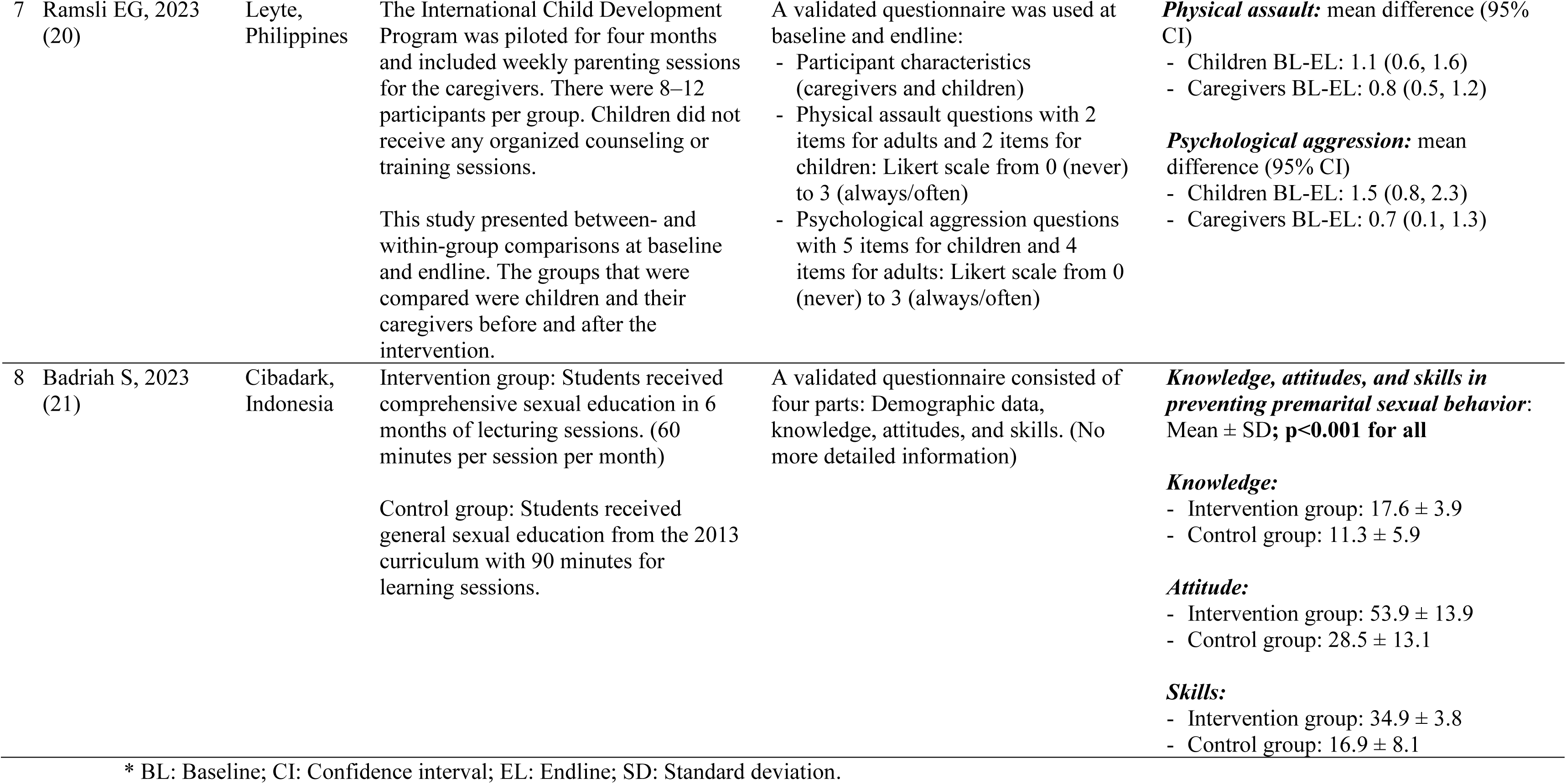
Characteristics and findings of included intervention studies.

### Study Quality Assessment

Based on the ROBINS-I tool for non-randomized control trials and the RoB-2 tool for randomized control trials (**Supplement Figure 2** for more detail) (13), the risk of bias in a cluster randomized controlled trial was the “some concerns” level (19) due to deviations from the intended intervention. Meanwhile, the risk of bias in the non-randomized studies (14–18, 20, 21) ranged from moderate to serious level. Most risks of bias in the non-randomized studies came from the biased selection of participants, bias in the classification of the intervention, and bias in the deviations from the intended interventions.

### Evidence of effectiveness

#### Sexual violence

Seven out of eight studies assessed associations (14–18, 21) or impact (19) of programs related to sexual violence. After the intervention, four studies (15, 17, 18, 21) reported increasing changes in knowledge, attitude, or skills toward sexual violence prevention between the interventional group and the control group, but there were no changes in the outcomes across the intervention and control groups in another study (14). One study (16) showed mixed results that positive changes in knowledge scores after watching the animated video were found in the 5th-grade students in Bali, but no difference after playing the game of snakes and ladders in this group. A cluster randomized trial (19) found no difference between the intervention and control schools for any outcomes such as perceived behavioral control, gender equality attitudes, socio-emotional learning competencies, descriptive norms, and intentions toward the sexual education program (data were not shown for the control group).

### Physical violence

In one pretest and post-test study (20), children reported a lower incidence of physical assault after the International Child Development Program Intervention. Specifically, children’s mean scores of physical assaults at baseline and end-line were 1.7 and 0.5, respectively (mean difference: 1.1; 95% CI: 0.6, 1.6). In contrast, one quasi-experimental study (14) found no difference in attitudes toward physical violence prevention between the intervention group and the control group after intervention [Intervention group (mean ± SD): 26.3 ± 2.9 and control group: 26.3 ± 2.6; p= 0.902].

#### Psychological violence

One pretest and post-test study found a lower incidence of psychological aggression after the International Child Development Program Intervention (20). Children’s mean scores of psychological aggressions at baseline and end-line were 2.2 and 0.7, respectively (mean difference: 1.5; 95% CI: 0.8, 2.3).

## Discussion

To our knowledge, this systematic review is the first to synthesize the findings from intervention studies designed to prevent various forms of GBV among adolescents in Southeast Asia. As such, this review provides a foundation for understanding the rigor, geographic scope, and outcomes of such studies in the region, and can guide future research and policy to prevent GBV among adolescents in Southeast Asia, who are known to be at high risk of exposure to GBV (2, 3).

Overall, our review identified very few intervention studies (n=8) designed to address GBV among adolescents aged 10 to 19 years in Southeast Asia. Moreover, the quality of the included studies was generally weak. Only one study was a randomized controlled trial. Two-thirds of the studies (n=5) had small sample sizes of less than 150, raising questions about the power to detect significant program effects. Most (n=7) studies did not directly measure behavioral GBV outcomes. Overall, the quality ratings were either “some concerns” or “moderate to serious” suggesting that improvements are needed throughout the region in the rigor of intervention studies to prevent GBV among adolescents. Finally, the geographic scope of intervention studies was extremely limited in Southeast Asia with most studies coming from Indonesia, so we are unable to generalize the findings from this review to all countries in the region. This review, instead, has identified a major regional gap in evidence to understand how GBV prevention intervention studies among adolescents aged 10 to 19 years may operate in Southeast Asia.

For the outcomes of included intervention studies designed to address GBV among adolescents, we found inconsistent results. Some studies found greater improvements in knowledge, attitudes, and skills related to GBV prevention in the intervention (versus control) group in the settings where studies were conducted in Indonesia and Thailand; however, other studies in these countries did not find any differences between the intervention and control groups. Future intervention studies need to assess program impacts on GBV behaviour.

Regarding ***sexual violence***, we found inconsistent findings across the included studies (14–18, 21). Some studies showed positive changes in knowledge, attitudes, or skills toward sexual violence prevention in the intervention group versus the control group (15, 17, 18, 21). Other studies showed no difference among these groups (14, 19). These conflicting results could be explained by the authors’ use of different data collection tools, limitations in the study design, and small sample sizes. A self-administrated questionnaire was used in two studies (14, 19) compared with a validated tool in other studies (17, 21). The inclusion of one cluster randomized controlled trial (19) and three quasi-experimental studies (15, 17, 21) also may have contributed to the mixed findings, since the latter studies may not have been able to account for confounding. The sample size was more than 465 participants in two studies (14, 19) but substantially smaller (< 150) in three studies (17, 18, 21), which most likely limited the power to detect significant pre-post differences between the intervention and control groups.

For ***physical violence***, we also found conflicting results (14, 20). After intervention, one pretest and post-test study in the Philippines (20) reported a lower incidence of physical assault, measured using a validated tool in 92 participants (46 children and 46 caregivers; female: 65%); whereas, one quasi-experimental study in Thailand found no evidence for a difference in attitudes about physical violence prevention using a self-administered questionnaire in 525 students (female: 47%) (14). However, it is hard to compare different physical violence outcomes within only two studies. Based on the paucity of data on intervention effects on attitudes and behavior toward physical violence, intervention studies among adolescents are needed throughout Southeast Asia to provide evidence-based guidance to policymakers about the best policies and programs to prevent physical violence among adolescents in the region.

Only one pretest and post-test study in the Philippines assessed ***psychological violence*** (20). This study demonstrated a lower incidence of psychological aggression at the endline versus baseline. This finding should be re-assessed in future RCTs measuring psychological violence outcomes among adolescents in Southeast Asia. Moreover, future research may consider specific forms of adolescent psychological violence such as threats and intimidation, denigration, ridicule, discrimination, rejection, and other non-physical forms of hostile treatment (22).

Taken together, very few studies in Southeast Asia have assessed associations or impacts of GBV interventions among adolescents aged 10 to 19 years, and evidence regarding the effectiveness of GBV intervention programs in this age group is generally weak. The conflicting results of included studies should be resolved with rigorous comparative RCTs in multiple countries in the region that assess primary behavioral outcomes related to all types of GBV and secondary outcomes related to knowledge, attitudes, and self-efficacy to change one’s behavior or intervene as a bystander. Based on the strongest evidence from this review, intervention programs that incorporate classroom activities and online learning with Facebook/Zalo/Instagram assignments and games may show promise and warrant more rigorous and widespread evaluation among adolescents in Southeast Asia.

## Conclusions

This systematic review revealed a paucity of intervention studies of limited quality to prevent GBV among adolescents in Southeast Asia. Conflicting findings were observed among included intervention studies for reasons that may relate in part to the low rigor of the intervention study designs. Interventions that integrate classroom activities with online learning via Facebook/Zalo/Instagram assignments and games may be appropriate for adolescents in this region. Rigorous RCTs across Southeast Asia are needed to expand the evidence base regarding what works to prevent GBV among adolescents in this region.

## List of abbreviations

CI: Confidence interval
GBV: Gender-based violence
GDP: Gross domestic product
IPV: Intimate partner violence
PRISMA: Preferred reporting items for systematic review and meta-analysis
RCTs: Randomized control trials

## Declarations

### Ethics approval and consent to participate

The National Institute for Health Research of the University of York approved the protocol registration for a systematic review (PROSPERO ID number: CRD42023476059). This systematic review does not contain any studies with human or animal subjects performed by any of the authors.

### Consent for publication

Not applicable

### Availability of data and materials

Not applicable

### Competing interests

Hoa H Nguyen, Kien G To, Van TH Hoang, Lu Gram, and Kathryn M Yount declare no competing interests.

## Funding

Not applicable

### Authors’ contributions

Writing protocol and register in the PROSPERO: All authors. Screening potential articles, data extraction and risk of bias assessment: Hoa H Nguyen and Kien G To. Results interpretation: All authors. Drafting manuscript: Hoa H Nguyen and Kathryn M Yount. Revising manuscript: All authors. Approval of final manuscript: All authors

## Data Availability

All data produced in the present work are contained in the manuscript

## Acknowledgments

We thank Mai NC Nguyen for screening potential articles and Mr. Glen Tittor for editorial assistance.

## Notes

### Competing Interest Statement

The authors have declared no competing interest.

### Funding Statement

This study did not receive any funding

